# Precipitating factors, presentation and outcomes of diabetic ketoacidosis among patients seen at moi teaching and referral hospital, eldoret kenya

**DOI:** 10.1101/2024.03.29.24304997

**Authors:** Clemence Mwahe Msagha, Jemima Kamano, Paul Ayuo

## Abstract

**Background:** Diabetes Ketoacidosis (DKA) is a major complication of Diabetes Mellitus (DM) with a likelihood of high mortality if not managed appropriately. It is diagnosed with a triad of hyperglycemia, ketonemia and metabolic acidosis.

**Objectives:** To describe the precipitating factors, clinical presentation and outcomes of DKA among patients attending Moi Teaching and Referral Hospital (MTRH).

**Methods:** This prospective study involved 120 consecutively recruited participants diagnosed with DKA. Participants were drawn from the Emergency department and Diabetes Outpatient clinic and followed up in the wards and intensive care unit (ICU) in MTRH for up to 10 days. Focused history and physical examination was done. Blood sugar was measured daily; blood ketones and blood gases were measured on days 1,2,3 and 5. Precipitating factors, presentation and outcomes were summarised as frequencies and their corresponding percentages and presented in tables and charts.

**Results:** The median age of participants was 33 years (IQR 23, 44.5). Type 1 DM represented 63.3% and type 2 DM 34.2% of the patients. The most common precipitating factors for DKA were; new onset undiagnosed DM (37.5%), missed medication (36.7%) and infection (35.8%). The most common presentation was dehydration (97.5%) with 49.2% of the patients having severe DKA while 22.5% had mild DKA. Urine and blood ketones for diagnosis of DKA were present in 46.4% and 100% of patients respectively. The median length of hospital stay was 6 days (IQR 5,7) with infection being a significant determinant (aOR 2.63). The number of days taken for DKA to resolve ranged from 1 to 5 days with a median period of 3 days (IQR 2,3). DKA in-hospital mortality was 9.2% with new onset DM being a significant determinant (uOR 5.19).

**Conclusion:** Some of the identified DKA precipitants in the study are preventable. The impact of DKA in MTRH is notable given the significant hospital stay and mortality.

**Recommendation:** We recommend implementation research studies that would develop and test different strategies to address the precipitants to prevent DKA. For the hospital to undertake an audit of current DKA management process with the aim of improving outcomes in terms of hospital stay and mortality.

## Introduction

Diabetes Mellitus (DM) is ranked by World Health Organization (WHO) as the fourth contributor to mortality due to non-communicable diseases (Alwan et al., 2000). Diabetes mellitus is characterized by chronic hyperglycaemia with disturbances of carbohydrate, fat and protein metabolism resulting from defects in insulin secretion, action or both (WHO, 2016).

There are various classification of diabetes mellitus but there are three main types: type 1 diabetes mellitus (T1DM) (10%), type 2 diabetes mellitus (T2DM) (85%), gestational diabetes mellitus (5%) and others (Brison, 2017).

The global prevalence of diabetes mellitus is gradually rising, especially in the low and middle income countries. The International Diabetes Federation (IDF) reported that the global prevalence of diabetes mellitus (DM) was 9.3%, translating to 463 million people affected by 2019 and this number is expected to increase to 10.2% translating to 578 million people (25% increment) by 2030, and 10.9% that is 700 million people (51% increment) by 2045 (International Diabetes Federation, 2019).

Currently on the recent updates of the International Diabetes Federation 2022 there are 537 million adults living with diabetes worldwide, 8.75 million individuals worldwide with T1DM and 528.25 million individuals with T2DM. In 2022, there were 530,000 new cases of T1DM diagnosed at all ages with 201,000 of these less than 20yrs of age. Sixty-two percent of all new T1DM cases were in people aged 20yrs or older (IDF, 2022).

A meta-analysis done by Peer et al and Azevedo et al looking at diabetes in Africa region involving countries like Kenya, Mali, Mozambique, Nigeria, South Africa and Zambia shows a surge in the condition, there were estimates of 39,000 people suffering from T1DM in 2013 with 6.4 new cases occurring per year per 100,000 people in children and adolescent <14 years old. The prevalence of T2DM among 20-79 year olds was (4.9%) with the majority of people with diabetes <60 years old; the highest proportion (43.2%) was in those aged 40-59 years. They reported a projected increase in numbers rising from 19.8 million in 2013 to 41.5 million in 2035, representing a 110% absolute increase. There was apparent increase in diabetes prevalence with economic development with rates of (4.4%) in low income countries, (5%) in lower middle income countries and (7%) in upper middle income countries (Peer, Kengne, Motala, & Mbanya, 2014)(Azevedo & Alla, 2008).

As per world health organization, the estimated prevalence of DM in Kenya was 3.3% in 2018 and predicted to reach 4.5% by 2025. This was backed up by the Kenya national STEPS survey which is a simple, standardized method for collecting, analysing and disseminating data on key non-communicable disease risk factors in countries comprising three steps. However, the study concluded that two thirds of the people living with diabetes may be undiagnosed and therefore not captured (Mensah, Korir, Nugent, & Hutchinson, 2020). Another study done in Kenya concluded that Kenya has an opportunity to reduce the burden of diabetes but funding needs to be concentrated on public health and primary healthcare interventions and people needing to adopt healthy lifestyles (Jones, 2013).

Diabetes Ketoacidosis (DKA) is a major life threatening complication of Diabetes Mellitus (DM) and is one of the most fatal acute complications of DM. It is characterized by hyperglycaemia, metabolic acidosis and ketonemia and occurs due to an absolute or relative deficiency of insulin in the body. Without enough insulin, the body begins to break down fat as fuel. This causes a build-up of acids in the blood called ketones and when left untreated the build-up causes DKA. DKA being an acute complication of diabetes, is associated with high rates of hospital admissions.

The prevalence of DKA in a study done on the youth in Colorado, DKA occurred in 25-30% of patients with T1DM and 4-29% of patients with T2DM (Dabelea et al., 2014).

The prevalence of DKA among hospitalised diabetes patients in a study in Hawassa Ethiopia was 40%, with 28.7% among type 1 DM patients and 11.28% among type 2 DM patients (Bedaso, Oltaye, Geja, & Ayalew, 2019). Another study done in Nigeria on the prevalence and clinical pattern of acute and chronic complications in African diabetes patients done by Jasper US et al, the prevalence of DKA was 12.2% (U.S. Jasper, n.d.). A study in Kenya at Kenyatta National Hospital (KNH) showed that DKA occurred in 8% of hospitalised diabetes patients (Mbugua, Otieno, Kayima, Amayo, & McLigeyo, 2005).

The diagnosis of DKA entails looking at the glucose concentration, presence of ketones and confirmation of acidosis. The use of blood ketones rather than urine ketones is advantageous because in DKA the first major ketone produced is 3 beta hydroxybutyrate (3-β-OHB) which is not detected by (urine sticks) urine acetoacetate (nitroprusside reaction). In DKA the 3-β-OHB: acetoacetate ratio increases from 1:1 to 5:1. With treatment of DKA, 3-β-OHB is oxidised back to acetoacetate. As a result, 3-β-OHB will decrease, but acetoacetate will increase. Measuring acetoacetate with urine sticks may therefore initially underestimate the severity of DKA and then continue to yield positive readings after the resolution of DKA. Another reason is that people with DKA are usually dehydrated at the time of presentation, and thus, urine output is reduced; it may take several hours after rehydration before urine is produced, further delaying the diagnosis and management (Brewster, Curtis, & Poole, 2017). Although blood gas analysis is typically run on arterial sample the United Kingdom (UK) guideline suggests the use of venous blood gas rather than arterial blood gas with a pH <7.3 for diagnosis of acidosis; because of the data suggesting that the differences between arterial and venous pH are not large enough to change clinical management decisions (Dhatariya & Vellanki, 2017a).

The most common precipitating factor in the development of these acute complications of diabetes mellitus, like Diabetic Ketoacidosis or Hyperosmolar Hyperglycaemic State (HHS) is infection (Kitabchi, Umpierrez, Miles, & Fisher, 2009). Other precipitating factors include major stresses that act by increasing insulin requirements and increasing secretion of the counter regulatory hormones including; myocardial infarction and cerebrovascular accident (CVA), alcohol abuse, pancreatitis, trauma, and drugs. In addition, new-onset T1DM or discontinuation of or inadequate insulin in established T1DM commonly leads to the development of DKA (Kitabchi et al., 2002). Drugs that affect carbohydrate metabolism, such as corticosteroids, thiazides, sympathomimetic agents (dobutamine and terbutaline) and cocaine may precipitate the development of HHS or DKA. (Alakkas, Alzaedi, Somannavar, & Alfaifi, 2020). (Warner, Greene, Buchsbaum, Cooper, & Robinson, 1998)

A study done by Polonsky et al showed that in young patients with type 1 DM, psychological problems complicated by eating disorders may be a contributing factor in (20%) of recurrent ketoacidosis. The factors that he found out that led to insulin omission in younger patients included fear of weight gain with improved metabolic control, fear of hypoglycaemia, rebellion from authority, and stress of chronic disease (Polonsky et al., 1994).

Although the symptoms of poorly controlled diabetes may be present for several days, the metabolic alterations typical of ketoacidosis usually evolve within a short time frame, typically <24 hr. DKA usually presents with history of polyuria, polydipsia and polyphagia because these patients have hyperglycaemia. They can also present with vomiting, abdominal pain, weakness, and because of the polyuria and vomiting they come in a dehydrated state, can have clouding of sensoria, and finally can present in a coma. Up to (25%) of DKA patients have vomiting, which may be coffee-ground in appearance. Mental status can vary from full alertness to profound lethargy or coma. In a study by Ahuja et al, the most common presentations were severe vomiting, abdominal pain and depressed mental state. (Ahuja, Kumar, Kumar, & Rizwan, 2019).Physical findings may include poor skin turgor and dry mucus membranes suggesting dehydration, Kussmaul respirations, tachycardia, hypotension, alteration in mental status, shock, and ultimately coma. Patients can be normothermic or even hypothermic primarily because of peripheral vasodilation unless there is an infection where they can be hyperthermic. Another cause of hypothermia is the lack of insulin preventing the intracellular movement of glucose to be used as a substrate for heat production. Hypothermia, if present, is a poor prognostic sign (Kitabchi et al., 2002).

The outcomes among patients admitted with DKA include: resolution of DKA, length of hospital stay and death (mortality). DKA mostly resolves in the first 48hrs when management has been well done following protocol (Trachtenbarg, 2005). Gosmanov et al reported that DKA can resolve in approximately 10hrs if strict management with protocols are observed (Gosmanov, Gosmanova, & Dillard-Cannon, 2014).

The mortality rate of DKA ranges from 2% to 5% in developed countries and 6% to 24% in developing countries. Without proper or appropriate management, it is 100% fatal (Bedaso et al., 2019). Several studies have found that the mortality rate of DKA has declined during the past 20 years.

A study in United States reported the mortality rate of DKA in developed countries as (<1%), but associating DKA with detrimental neurocognitive outcomes and (>5%) in the elderly patients and in those with life threatening conditions (Duca, Wang, Rewers, & Rewers, 2017), (Kitabchi et al., 2002). With the increasing health care costs and a changing healthcare system, prevention of diabetic ketoacidosis remains very essential.

A study in Tanzania reported a mortality rate of (24.1%) with a major cause of morbidity and mortality being admission to intensive care unit (ICU) (Iddi et al., 2017).

Mbugua et al described a mortality rate of (29.8%) within 2 days of admission with DKA in Kenyatta National Hospital (KNH) in Kenya (Mbugua et al., 2005).

In the adult population, hypokalaemia, adult respiratory distress syndrome/acute lung injury and co-morbid states such as pneumonia, acute myocardial infarction, renal failure and sepsis are all associated with increased mortality in DKA (Dhatariya, Glaser, Codner, & Umpierrez, 2020).

Moi teaching and referral hospital (MTRH) being the only level 6 facility in the western region of Kenya, receives both walk in patients with diabetes and referrals from the region and hence has a representative spectrum of DKA patients.

Therefore, this study was designed to determine the precipitating factors, clinical presentations and the outcomes of patients with diabetes admitted with DKA in MTRH.

## Materials and Methods

### Study Design

This study was a prospective study involving 120 patients with a diagnosis of DKA.

### Study Setting

The study was carried out at Moi Teaching and referral hospital (MTRH), the second national referral hospital in Kenya after Kenyatta National Hospital (KNH), located in Uasin Gishu County in the North Rift region. It serves as a referral hospital providing specialized care to clients in Western Kenya, Eastern Uganda and South Sudan with a catchment population of approximately 24 million.

### Target Population

Patients with diabetes presenting at MTRH accident and emergency unit and DM clinics. Inclusion criteria:

- Known and newly diagnosed DM patients who met DKA criteria
- Participants 13 years and above.

#### Study period

1 year (May 2021-May 2022) to achieve sample size. Participants were followed up inpatient to a maximum of 10 days.

#### Sampling technique

Consecutive recruitment of patients with diabetes presenting with diabetic ketoacidosis in the casualty, in DM clinic and in the wards was done.

### Sample size calculation

For objective one and two the sample size calculation was derived from the prevalence of DKA in KNH.

Mbugua et al, 2005 found out that DKA occurred in 8% of the hospitalized patients with DM in KNH. With the 95% confidence interval, Cochran formulae was used to determine the sample size.

Where, n = estimated minimal sample size in infinite population

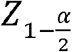 - is the 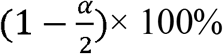 quartile of the standard normal distribution with type I error of α = 5%.

P – is the average prevalence of DKA from the previous study.

δ – is the margin of error taken to be 5 %.

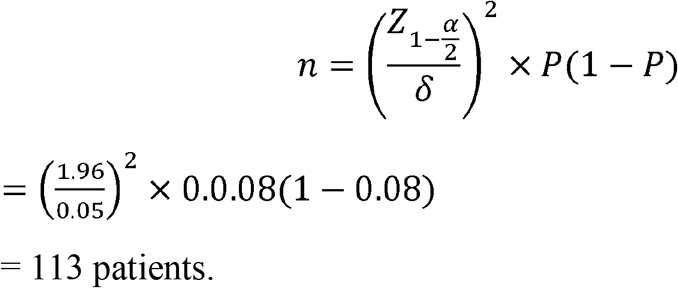

For objective three, the sample size calculation used was described by Kelsey et al for estimating a single population mean.

The sample size calculation was derived from a study on the treatment outcome of DKA among patients attending a general hospital in north-west Ethiopia (Mekonnen, Gelaye, Gebreyohannes, & Abegaz, 2022). It had a mean length of hospital stay of 4.6 days and the standard deviation of length of hospital stay was ±2.8 days. Out of the three outcomes: time to DKA resolution, length of hospital stay and mortality. We used the outcome of length of hospital stay because it was giving us the highest number of the sample size.

SD=±2.8, Precision=0.5

Where, n = estimated minimal sample size

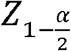 - is the 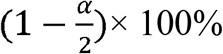 quartile of the standard normal distribution with type I error of α = 5%.

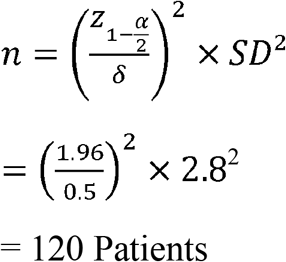

### Study Procedure/Data collection methods

Principal Investigator (PI) and research assistants screened for potential participants for eligibility, patients with known DM or with RBS>11.1mmol/l in the accident and emergency and diabetic clinics. Once participants were thought to be potential candidates for the study, the PI explained the study to them and participants were given consent forms in a language they could understand. After consenting, a pin prick was done to evaluate for blood ketones on the participants by the principal investigator using a blood ketone meter. Those with blood ketones <3mmol were excluded from the study but continued in care, while those with blood ketones >3mmol continued in the study and a blood sample for BGA was taken from them by the PI. Those found not to have acidosis were excluded from the study but continued care while those found to have acidosis continued in the study. DKA was confirmed by hyperglycemia, ketonemia and acidosis. Participants were followed up to the time to DKA resolution and discharge with a maximum of 10days follow-up for the inpatient. Those who were still hospitalized beyond day 10 were taken care of by the primary team. For the outpatient participants not requiring admission with mild DKA they were followed up to the point of resolution of DKA/discharge which was within 24hrs. Data on sociodemographic, precipitating factors and presentations was collected as per the data collection forms, which were interviewer administered. Most of the data captured in a structured questionnaire. Tests done during follow up were random blood sugar, blood ketones and blood gas analysis (BGA) to see the improvement of the patient up to the point of resolution of DKA. The random blood sugars were being done daily while the blood ketones and blood gas analysis were done on day 1, 2, 3 and 5. Patients were considered to be out of DKA when the blood ketones were <1mmol/L, and HCO3 >18mEq/L and PH >7.3.

### Data analysis

Data were analyzed using STATA, version 16 (StataCorp, College Station, Texas 77845 USA). Preliminary analysis involved summary of social demographic and clinical characteristics where categorical variables such as gender, marital status, educational level, employment status, types of diabetes and diagnosis status and follow up status as outpatient were summarized using frequencies and their corresponding proportions.

Numerical variables such as age, age at diabetes diagnosis, duration with diabetes, duration of follow-up were summarized as means and their corresponding standard deviations if they assumed normal distribution else were summarized as median and their interquartile ranges.

#### Objective 1 and 2

Different Presentations and Precipitating factors were summarised as frequency and respective percentages then were presented in form of tables.

#### Objective 3

The length of stay was summarised using median and interquartile and later grouped into two categories 1-5 days and 6-10days and summarized as frequency and percentage.

Other outcomes like, resolution of DKA and death were summarised as frequency and their corresponding percentages.

A sub analysis was done to determine the association between study outcomes and patient’s characteristics using logistic regression. This was an exploratory analysis based on the findings and since it was not part of my objectives it was not powered enough but we saw it as a priority to mention.

The findings were presented in form of tables, charts. All the test results were considered statistically significant if p-value was less than 0.05.

### Ethical Consideration

Ethical clearance was obtained from Moi University Institutional Research Ethics Committee (IREC) before the conduct of the research, IREC no. FAN:0003866. Permission to conduct the study was sought from MTRH management and National Commission for Science, Technology and Innovation (NACOSTI) NACOSTI/P/21/11944.

Written informed consent was sought from the participants before enrolment in the study, and they were allowed to withdraw from the study if they chose to at any point. For the patients below 18yrs of age assent was sought from them and consent was sought from the parent/guardian or immediate caretakers. For patients who were unable to consent because of reduced level of consciousness consent was sought from the relative.

This study involved no intervention on the participants and the procedures the patients underwent like history taking, physical examination and blood samples drawn are usually done routinely during care and consent to participate in the study included consent for blood draws. Therefore, no harm was anticipated.

There was no monetary incentives or benefits to the participants.

The results were duly communicated to the primary physician for action to be done and confidentiality was maintained throughout the whole study period. Numerical codes were used to number the questionnaires and as identity to the study subjects so as to maintain confidentiality. Stored data was encrypted with the password only known to the Principal investigator. The filled questionnaires, signed consent forms and other data collection tools were kept in a locked place in the custody of the Principal investigator.

## Results

### Sociodemographic characteristics

The age of patients that presented in DKA ranged from 14 to 89 years with a median age of 33 (IQR 23, 44.5) years. While the age at diabetes diagnosis ranged from 7 to 75 years with a median age of 30 (IQR 20, 40.5). The duration with diabetes ranged from 3months to 33 years for those known to have diabetes with median duration of 5(IQR 2, 8) years. The rest on the attached table.

**Table 1:** Sociodemographic characteristics.

| **Characteristic** | **Frequency** | **Percentage** |
| --- | --- | --- |
| **Age** |  |  |
| **Mean (SD)** | 35(15) |  |
| **Range** | 14-89 |  |
| **Gender** |  |  |
|  | **n 120** |  |
| Male | 52 | 43.3 |
| Female | 68 | 56.7 |
| **Marital status** |  |  |
|  | **n 120** |  |
| Married | 56 | 46.7 |
| Single | 46 | 38.3 |
| Separated | 12 | 10.0 |
| Widowed | 6 | 5.0 |
| **Education level** |  |  |
|  | **n 120** |  |
| None | 4 | 3.3 |
| Primary | 55 | 45.8 |
| Secondary | 42 | 35.0 |
| Tertiary | 19 | 15.8 |
| **Employment status** |  |  |
|  | **n 120** |  |
| Formally employed | 11 | 9.2 |
| Self-employed | 22 | 18.3 |
| Unemployed | 87 | 72.5 |
| **Living arrangement** |  |  |
|  | **n 120** |  |
| Alone | 12 | 10.0 |
| With others | 108 | 90.0 |

**Table 2:** Clinical characteristics.

| **Characteristic** | **Frequency** | **Percentage** |
| --- | --- | --- |
| **Diabetes type** | **n 120** |  |
| I | 76 | 63.3 |
| II | 41 | 34.2 |
| Gestational | 1 | 0.8 |
| Secondary | 2 | 1.7 |
| **Diagnosis status** | **n 120** |  |
| Newly diagnosed DM on admission | 45 | 37.5 |
| Known diabetic | 75 | 62.5 |
| **Follow-up status as outpatient** | **n 75** |  |
| On follow-up | 50 | 66.7 |
| Lost to follow-up | 19 | 25.3 |
| No follow-up | 6 | 8 |
| **Current medication as outpatient** | **n 120** |  |
| Insulin | 58 | 48.33 |
| Oral glucose lowering agent (OGLA) | 14 | 11.67 |
| Insulin & OGLA | 3 | 2.50 |
| Other drugs (hypertensive drugs) | 5 | 4.17 |
| None | 40 | 33.33 |
| **Medication adherence** | **n 75** |  |
| No | 47 | 62.7 |
| Yes | 28 | 37.3 |
| **Reason for non-adherence** | **n 47** |  |
| Financial constraints | 22 | 46.81 |
| Forgets | 11 | 23.40 |
| Ran out of drugs | 2 | 4.26 |
| Travels a lot | 3 | 6.38 |
| Couldn't explain | 1 | 2.13 |
| Defaulted felt stable | 4 | 8.51 |
| Lipohypertrophy | 1 | 2.13 |
| Not been feeding well on chemo | 1 | 2.13 |
| Substance abuse | 1 | 2.13 |
| Uses herbal meds | 1 | 2.13 |
| **Admission in last one year** | **n 120** |  |
| No | 66 | 55 |
| Yes | 54 | 45 |

**Table 3:** Precipitating factors.

| **Precipitating factors** | **Frequency(n120)** | **Percentage** |
| --- | --- | --- |
| Newly onset DM | 45 | 37.5 |
| Missed doses (non-adherence) | 44 | 36.7 |
| Infection | 40 | 35.8 |
| Stressful conditions | 15 | 12.5 |
| Under dosing and Inappropriate feeding | 11 | 9.2 |
| Poor insulin storage | 4 | 3.3 |
| Infection with missed doses | 9 | 7.5 |
| Infection with new onset | 5 | 4.2 |

**Table 4:** Types of infections recorded.

| **Precipitating factors** | **Frequency (n120)** | **Percentage** |
| --- | --- | --- |
| Pneumonia | 18 | 15 |
| UTI | 9 | 7.5 |
| TB | 3 | 2.5 |
| Septic diabetic foot | 2 | 1.7 |
| Meningoencephalitis | 2 | 1.7 |
| Cellulitis | 2 | 1.7 |
| Otitis media | 2 | 1.7 |
| Catheter site infection | 2 | 1.7 |

**Table 5:** Clinical presentation.

| **Presentation** | **Frequency (n120)** | **Percentage** |
| --- | --- | --- |
| Polydypsia | 114 | 95 |
| Polyuria | 112 | 93.3 |
| General weakness | 102 | 85 |
| Nausea/Vomiting | 77 | 64.2 |
| Abdominal pain | 77 | 64.2 |
| Difficulty in breathing | 64 | 53.3 |
| Weight loss | 57 | 47.5 |
| Polyphagia | 55 | 45.8 |
| Others | 44 | 23.9 |
| **Type of DKA** | **Frequency** | **Percentage** |
| Euglycemia | 4 | 3.3 |
| Mild | 27 | 22.5 |
| Moderate | 30 | 25.0 |
| Severe | 59 | 49.2 |
| **GCS** | **Frequency** | **Percentage** |
| Normal (15/15) | 55 | 45.8 |
| Abnormal ($\leq 14/15$) | 65 | 54.2 |
| **Hydrationstatus (Dehydrated)** |  |  |
| No dehydration | 3 | 2.5 |
| + | 3 | 2.5 |
| ++ | 114 | 95 |
| **Kussmaul breathing** |  |  |
| Absent | 40 | 33.3 |
| Present | 80 | 66.7 |

**Table 6:**
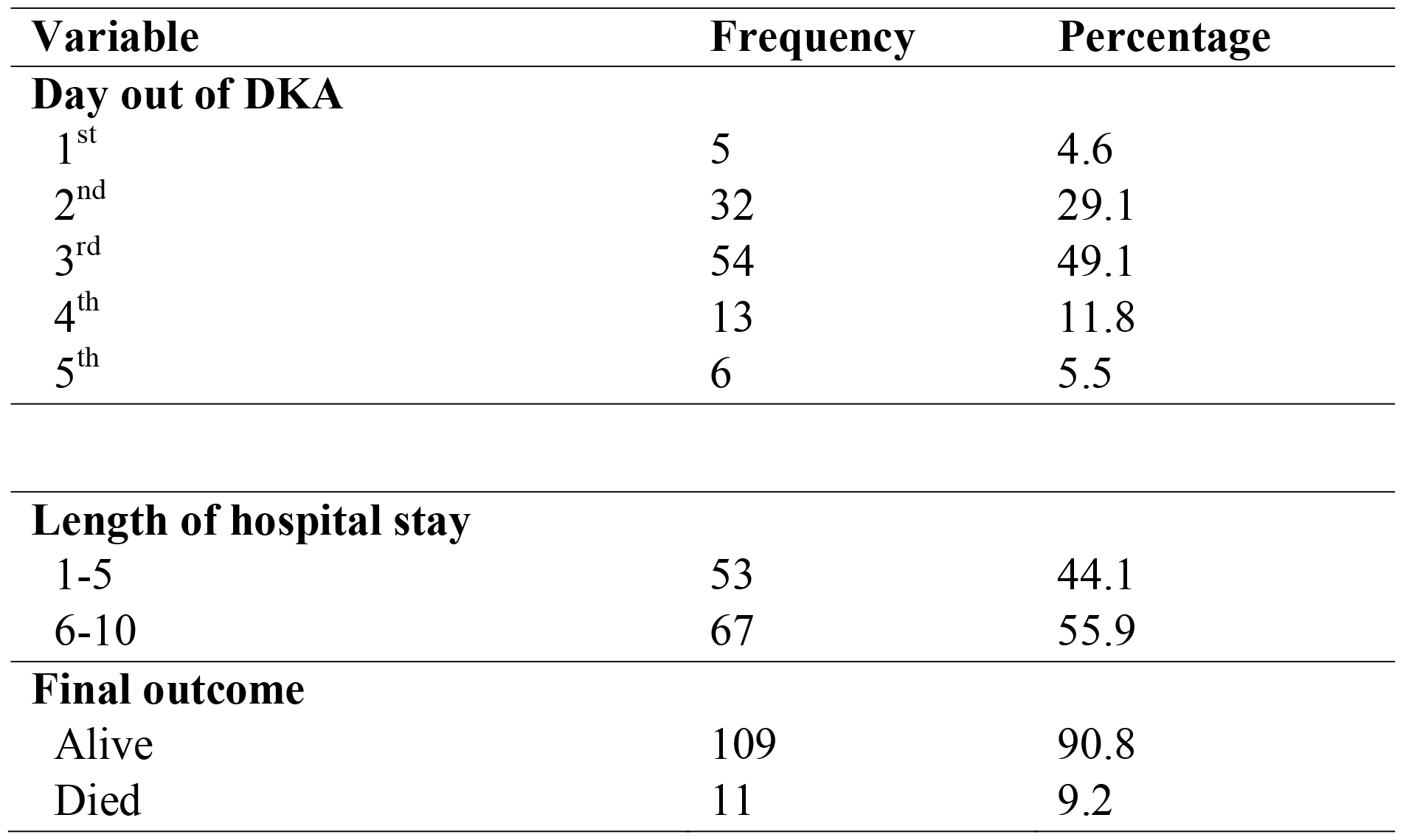
In-hospital outcomes.

**Table 7:** Association of length of hospital stay and patient’s characteristics.

| Characteristic | Follow-up days |  | uOR | 95% CI | p-value | aOR | 95% CI | p-value |
| --- | --- | --- | --- | --- | --- | --- | --- | --- |
|  | 1-5 days | 6-10 days |  |  |  |  |  |  |
| Age |  |  |  |  |  |  |  |  |
| Median (IQR) | 31 (21, 44) | 35 (26, 45) | 1.009 | 0.98-1.03 | 0.471 |  |  |  |
| Sex |  |  |  |  |  |  |  |  |
| Male | 26(50) | 26(50) | Ref |  |  |  |  |  |
| Female | 27(39.7) | 41(60.3) | 1.52 | 0.73-3.15 | 0.261 |  |  |  |
| DM type |  |  |  |  |  |  |  |  |
| Type I | 35 (46.1) | 41(53.9) | Ref |  |  |  |  |  |
| Type II | 17(41.5) | 24(58.5) | 1.21 | 0.56-2.60 | 0.634 |  |  |  |
| Others | 1(33.3) | 2(66.7) | 1.71 | 0.15-19.64 | 0.668 |  |  |  |
| Diagnosis duration |  |  |  |  |  |  |  |  |
| Newly diagnosed DM | 25(55.6) | 20(44.4) | Ref |  |  |  |  |  |
| Known DM patient | 28(37.3%) | 47(62.7) | 2.10 | 0.99-4.45 | 0.053 |  |  |  |
| **Admitted in the last 1 year** |  |  |  |  |  |  |  |  |
| No | **41(62.1)** | **25(37.9)** | **Ref** |  |  | **Ref** |  |  |
| Yes | **12(22.2)** | **42(77.9)** | **5.74** | **2.55-12.93** | **<0.001** | **5.91** | **2.57-13.59** | **<0.001** |
| DKA type |  |  |  |  |  |  |  |  |
| Mild | 15(55.6) | 12(44.4) | Ref |  |  |  |  |  |
| Moderate | 17(56.7) | 13(43.3) | 0.96 | 0.02-2.74 | 0.933 |  |  |  |
| Severe | 20(33.9) | 39(66.1) | 2.44 | 0.96-6.18 | 0.061 |  |  |  |
| **Having infection** |  |  |  |  |  |  |  |  |
| No | **40(51.9)** | **37(48.1)** | **Ref** |  |  | **Ref** |  |  |
| Yes | **13(30.2)** | **30(69.8)** | **2.49** | **1.13-5.49** | **0.023** | **2.63** | **1.11-6.23** | **0.027** |

**Table 8:** Association of death and patient’s characteristics.

| Characteristic | Final outcome |  | uOR | 95% CI | p-value |
| --- | --- | --- | --- | --- | --- |
|  | Alive | Died |  |  |  |
| Age |  |  |  |  |  |
| Median (IQR) | 33(24, 45) | 28(17, 44) | 0.98 | 0.94-1.02 | 0.411 |
| Sex |  |  |  |  |  |
| Male | 46(88.5) | 6(11.5) | Ref |  |  |
| Female | 63(92.6) | 5(7.4) | 0.61 | 0.17-2.12 | 0.435 |
| DM type |  |  |  |  |  |
| Type I | 68(89.5) | 8(10.5) | Ref |  |  |
| Type II | 38(92.7) | 3(7.3) | 0.67 | 0.16-2.68 | 0.572 |
| Others | 3(100) | 0 | NA | NA |  |
| Diagnosis duration |  |  |  |  |  |
| **Known DM patient** | **72(96)** | **3(4)** | **Ref** |  |  |
| **Newly diagnosed DM** | **37(82.2)** | **8(17.8)** | **5.19** | **1.30-20.73** | **0.020** |
| Admitted in the last 1 year |  |  |  |  |  |
| No | 59(89.4) | 7(10.6) | Ref |  |  |
| Yes | 50(92.6) | 4(7.4) | 0.67 | 0.18-2.43 | 0.548 |
| DKA type |  |  |  |  |  |
| Moderate | 27(90) | 3(10) | Ref |  |  |
| Mild | 27(100) | 0 | NA | NA |  |
| Severe | 51(86.4) | 8(13.6) | 1.41 | 0.35-5.76 | 0.631 |
| Having infection |  |  |  |  |  |
| No | 70(90.9) | 7(9.1) | Ref |  |  |
| Yes | 39(90.7) | 4(9.3) | 1.03 | 0.28-3.72 | 0.969 |

**Table 9:** RBS findings over time.

| **Days** | **RBS Category** |  |  |
| --- | --- | --- | --- |
|  | **Normal** | **High** | **Very high** |
| **1 (n=120)** | 1(0.8) | 12(10) | 107(89.2) |
| **2 (n=109)** | 4 (3.7) | 53(48.6) | 52(47.8) |
| **3 (n=109)** | 22(20.2) | 75(68.8) | 12(11) |
| **4 (n=107)** | 43(40.2) | 64(59.8) |  |
| **5 (n=101)** | 77(76.2) | 24(23.8) |  |
| **6 (n=67)** | 58(86.6) | 9(13.4) |  |
| **7 (n=43)** | 39(90.7) | 4(9.3) |  |
| **8 (n=17)** | 14(82.3) | 3(17.7) |  |
| **9 (n=16)** | 14(87.5) | 2(12.5) |  |
| **10 (n=16)** | 16(100) |  |  |

**Table 10:** Blood ketones findings over time.

| **Serum ketones category** |
| --- |

| **Days** | **Normal** | **High** | **Very high** | **Extremely high** |
| --- | --- | --- | --- | --- |
| **1 (n=120)** |  | 70 (58.3) | 25 (20.8) | 25 (20.8) |
| **2 (n=72)** | 54(75) | 12(16.7) | 6(8.3) |  |
| **3 (n=89)** | 73(82) | 13(14.6) | 3(3.4) |  |
| **5 (n=11)** | 10(90.9) |  | 1(9.1) |  |

**Table 11:** BGA findings from day 1-5.

| **BGA Category** | **Days** |  |  |  |  |
| --- | --- | --- | --- | --- | --- |
|  | **Day 1** | **Day 2** | **Day 3** | **Day 4** | **Day 5** |
| **PH** |  |  |  |  |  |
| Normal(7.35-7.45) | - | 17 (17.3) | 40(45.5) | 20(64.5) | 11(100) |
| Low (7.0-7.34) | 38(31.7) | 45 (45.9) | 30(34.1) | 11(35.5) |  |
| Very low(<7.0) | 82(68.3) | 36 (36.7) | 18(20.5) |  |  |
| **HCO3** |  |  |  |  |  |
| Normal(22-26) | - | 4 (4.1) | 34(38.6) | 20(64.5) | 11(100) |
| Low(15-21) | 14(11.6) | 52 (53.1) | 42(47.7) | 11(35.5) |  |
| Very low(<15) | 106(88.3) | 42 (42.9) | 12(13.6) |  |  |
| **AG** |  |  |  |  |  |
| Normal (12-16) | 21 (17.5) | 32 (32.6) | 38(43.2) | 20(64.5) | 11(100) |
| High (>16) | 99 (82.5) | 66 (67.3) | 50(56.8) | 11(35.5) |  |

**Table 12:** Other laboratory findings on day 1.

| **Characteristic** | **Frequency** | **Percentage** |
| --- | --- | --- |
| **Urinalysis** |  |  |
| **Urine ketones (n=112)** |  |  |
| Negative | 29 | 25.9 |
| + | 31 | 27.7 |
| ++ | 23 | 20.5 |
| +++ | 29 | 25.9 |
| **Urine glucose (n=111)** |  |  |
| + | 8 | 7.2 |
| ++ | 26 | 23.4 |
| +++ | 73 | 65.8 |
| ++++ | 3 | 2.7 |
| Anuric | 1 | 0.9 |
| **Urine protein (n=47)** |  |  |
| + | 24 | 51.1 |
| ++ | 12 | 25.5 |
| +++ | 1 | 2.1 |
| Trace | 10 | 21.3 |
| **Blood (n=35)** |  |  |
| + | 13 | 37.1 |
| ++ | 9 | 25.7 |
| +++ | 13 | 37.1 |
| **Nitrite (n=7)** |  |  |
| + | 7 | 100 |
| **Leucocytes (n=6)** |  |  |
| + | 1 | 16.7 |
| ++ | 3 | 50 |
| +++ | 2 | 33.3 |
| **Blood samples** |  |  |
| **PCT (n=28)** |  |  |
| Normal <0.1ng/ml | 9 | 32.1 |
| High >2 | 19 | 67.9 |
| **WBC (n=120)** |  |  |
| Low (<4) 10<sup>9</sup>/L | 4 | 3.3 |
| Normal (4-10) | 40 | 33.3 |
| High (>10) | 76 | 63.3 |
| **Absolute Neutrophil count (n=120)** |  |  |
| Low (<2) 10<sup>9</sup>/L | 4 | 3.3 |
| Normal (2-7) | 43 | 35.8 |
| High (>7) | 73 | 60.8 |
| **CRP (n=120)** |  |  |
| Normal (0-5) mg/L | 65 | 54.7 |
| High (>5) | 55 | 45.8 |
| **HbA1c (n=116)** |  |  |
| High (>7.5%) | 116 | 100 |
| **UECs** |  |  |
| **Urea (n=120)** |  |  |
| Normal (0-8.3)mmol/L | 74 | 61.7 |
| High (>8.3) | 46 | 38.3 |
| **Creatinine (n=120)** |  |  |
| Low (<44)umol/L | 7 | 5.8 |
| Normal (44-80) | 58 | 48.3 |
| High (>80) | 55 | 45.8 |
| **Na (n=100)** |  |  |
| Low (<135)mmol/L | 66 | 66 |
| Normal (135-145) | 24 | 24 |
| High (>145) | 10 | 10 |
| **K (n=100)** |  |  |
| Low (<3.5)mmol/L | 12 | 12 |
| Normal (3.5-4.5) | 58 | 58 |
| High (>5.5) | 30 | 30 |

## Discussion

To the best of our knowledge this is a first longitudinal study looking at DKA precipitants, presentations and outcomes in western Kenya and hence provides very useful data on the condition. Despite concerted efforts to improve diabetes care in MTRH and any other facility, DKA still remains a common complication with significant morbidity and mortality.

The study was looking at patients presenting with DKA in both type 1 DM and type 2 DM, focusing on the precipitating factors, presentations and outcomes.

### Factors precipitating DKA among patients at MTRH

The study included a wide range of patients from 14-89 years of age with a mean age of 35±15yrs and the age of diagnosis of diabetes ranging from 7-75yrs. Indeed, this allowed representation of both T1DM and T2DM patients and emphasizes the fact that DKA affects both types of diabetes. T1DM of the patients presenting in DKA represented 63.3% whereas 34.2% were T2DM. It reflects findings of a study done in Texas which found out that 53% of the patients with DKA were T1DM, 39% were T2DM and 8% were not ‘typed’(Balasubramanyam et al., 1999).

The most common precipitating factor in this study was new diagnosis of diabetes mellitus with DKA as the primary presentation of DM at 37.5%. This is because most of the patients were T1DM. This was closely followed by missed doses (36.7%), then infections (35.8%). This was similar to a study in Ethiopia that had new onset diagnosis of DM at 38.8% and a study in Kenya at KNH by Mbugua et al where half of the patients studied were newly diagnosed (Mekonnen et al., 2022),(Mbugua et al., 2005). It was different from other studies that were getting infection as the most common precipitating factor (Seth et al., 2015) and others poor compliance/missed doses as the most common precipitating factor (Thewjitcharoen et al., 2019). The difference with the other studies is that they had a predominance of T2DM. Some of the patients had more than one precipitating factor, for example, some of the new onset diabetes were also having an infection at 4.2% and others had both infections and missed doses at 7.5%. The overlapping factors is supported by a study done in India which had (53%) of patients having both poor compliance and infection (Sonwani, Arya, & Saxena, 2018).

Most of the first presentation of diabetes (newly diagnosed DM) was associated with the younger age group as expected given the peak age of type 1 DM presentation. It can also be said that patients presenting newly with hyperglycaemia should be screened for DKA and other precipitating factors because they tend to overlap. This was similar to a study by Ahuja et al that had a statistical significance of younger age group associated with first presentation (Ahuja et al., 2019).

Among those who missed doses, some lacked insulin, others were taking lower dose than prescribed and this was teased out from history because they didn’t want the drug to finish very fast and their sugars were not well controlled with the dose they were using while others were skipping doses. Of note is that most of the participants had a primary level education (45.8%) and most of them were unemployed (72.5%) despite majority being 18years and above. This could explain the fact that financial constraint was a major concern that led to non-compliance of these patients. This is similar to the study in India where poor socioeconomic status resulted in patients being not compliant and having poor glycaemic control so any precipitating factor landed them to DKA (Sonwani et al., 2018).

Therefore, it can be explained that lack of adherence to treatment is an important precipitating factor to DKA and regular assessment for adherence is needed especially when reviewing patients in the DM clinics. DKA screening in non-adherent patients may also be a good strategy to reduce the occurrence of DKA. Of note this study found that (46.7%) of the participants were married, (38.3%) were single, (10%) were separated and (5%) were widowed. Ninety percent of these participants were living with others as the living arrangement while ten percent were living alone. This indicated that most of the participants had a support system and that there is potential to include the social support in the processes of care, and also educate them about diabetes and DKA and that may help in treatment support and potentially better treatment adherence.

The most common infection in this study was pneumonia (13.3%), followed by UTI (5.8%), then PTB (2.5%) a fact that may be useful in informing regular screening and also research on ways of preventing/diagnosing these infections early to prevent complications. Infection in this study was supported by procalcitonin (PCT) a biomarker specific for bacterial infections, CXR for pneumonia and PTB; Urinalysis for UTI and some blood cultures depending on the foci especially for those that had catheter related sepsis. We used a much more sensitive method for assessing infections that is why we had a bigger number than the rest of the studies.

This was closely related to a study in India which concluded that up to 50% of the patients had infection as a precipitating factor with the main infection being lobar pneumonia at 19.2%, pulmonary tuberculosis (PTB) 15.5% and urinary tract infection (UTI) at 9.6% (Chaudhary et al., 2016a).

Other interesting factors were CKD/DM patients who were off insulin and patients with malignancies that came in with euglycemic DKA, dialysis in the CKD patients helped in their management, this is similar to a case study that looked at DKA in a CKD patient (Varma & Karim, 2016).

### Clinical presentation of DKA in patients at MTRH

As expected the most common presentation in this study was a history of polydipsia (95%) and polyuria (93.3%) with general body weakness (85%) therefore these symptoms are still important to look for in patients presenting with DKA, other symptoms included nausea and vomiting (64.2%) with abdominal pain and weight loss but polyphagia was less common at 45.8%. Surprisingly most patients did not have appetite to eat at the time of presentation. This was different from a study in Ethiopia which found out that 50.3% of the patients with DKA had 3Ps (polydipsia, polyuria and polyphagia), 29.2% had 2 Ps (polyuria and polydipsia), 1.5% had weight loss while other signs and symptoms contributed 15.9% (Bedaso et al., 2019).

Not surprising, dehydration was the most common sign at the time of presentation of these patients (97.5%), most of them in moderate to severe dehydration. Dehydration was demonstrated by dry mucous membranes, skin turgor of > 2 seconds, increased heartrate and increased respiratory rate; for severe dehydration they additionally had low blood pressure and faint pulse. These were signs on physical examination. A point of care ultrasound was not done to check the inferior venacava (IVC) to evaluate for dehydration. The state of patients coming in a dehydrated state could suggest delayed health seeking behaviour in these patients. This was similar to a study by Chaudhary et al where most patients were presenting in a dehydrated state (Chaudhary, Singh, & Nigam, 2016b). Because patients were presenting in a dehydrated state it was difficult to get a urine sample for some of the patients at the time of presentation unless after hydration and one chronic kidney disease patient was completely anuric.

Most patients presented with Kussmaul breathing indicating the degree of acidosis, although (54.2%) of the patients had altered mental state (low GCS), none was in a coma and (45.8%) of the patients were fully alert at the time of presentation hence the need to do thorough screening for DKA even in DM patients who may not appear too sick. These findings were almost similar with a study done in India whose findings showed vomiting and abdominal pain in (34.9%) of patients, altered sensorium in (47%), Kussmaul breathing in (28%) and hypotension in (46%) of patients with DKA (Adhikari et al., 1997).

Tachycardia was present in (63.3%) of the patients and (53.3%) had a high respiratory rate at the time of presentation. This would have been contributed by the hyperglycaemia and the dehydration the patients presented with, and the Kussmaul breathing respectively. There was no documentation of any cardiac arrhythmias in any of the patients. This is closely related to a study in Saudi Arabia which showed tachycardia as the most common clinical sign noted in the patients on admission that presented with DKA (Almalki, Buhary, Khan, Almaghamsi, & Alshahrani, 2016).

Almost half (49.2%) of the patients had severe DKA with (25%) having moderate DKA and (22.5%) having mild DKA. The severity was determined mostly by a blood gas analysis looking at the pH, the HCO3 and the AG. Despite having patients with severe DKA, some had mild DKA and this emphasizes the matter of screening of DKA in stable patients while reviewing them in DM clinics. This was similar to a study that reviewed most of their participants to have moderate to severe DKA (Newton & Raskin, 2004a). It was different from a study by Mekonnen et al that had a predominance of mild DKA at 75.5% (Mekonnen et al., 2022).

Most of the patients that presented with DKA in this study were type 1 DM patients (63.3%), with type 2 DM being (34.2%) and (1.7%) having secondary diabetes. This could be explained by the fact that most of the patients presenting with DKA were of younger age or were already known T1DM. The type 1 DM predominance was similar to a study looking at the clinical and biochemical differences in type 1 DM and type 2 DM presenting with DKA (Newton & Raskin, 2004a) this was closely related to my study because they were cutting across different age groups. Another study done in England looking at diabetes in the youth found that DKA was more in type 1 DM than in type 2 DM (Zhong, Juhaeri, & Mayer-Davis, 2018) this was because it was focusing on the youth. It was different from other studies done in India and Thailand that documented majority of DKA patients being in type 2 DM rather than type 1 DM (Seth et al., 2015), (Thewjitcharoen et al., 2019), (Shahid et al., 2020) this was attributed to the fact that the prevalence of type 2 DM is much higher than type 1 DM.

The patients that presented with secondary diabetes in this study had a history of long term use of steroids that precipitated DKA as part of new onset diabetes mellitus. This has previously been described and it’s a known risk factor (Cavataio & Packer, 2022).

Our study recorded that most of the patients presenting in DKA were females (56.7%) than males (43.3%) which is consistent with the prevalence of DKA being more in females and the reason is unclear. It is in keeping with the MTRH diabetes clinic attendance proportions due to differential health seeking behaviour since (60%) of the patients in the clinics are females. It can also be explained by the hormonal changes in women due to their oestrogen levels. In addition, cytokines like IL 1, IL 6 and TNF produced during stress which is more common in women antagonises the effect of insulin leading to DKA. This female predominance is similar to a study in Thailand which had a female predominance of (61.5%) and males at 38.5% (Thewjitcharoen et al., 2019) and a study in India with a female predominance of 51.9 % (Chaudhary et al., 2016b). It was different from a study done in our neighbouring country in Mwanza Tanzania that had a male predominance (62.1%) compared to females (37.9%) but this was because their age group ranged from 1 year to > 30 years (Iddi et al., 2017).

Majority of the patients in our study (62.5%) presenting with DKA were those with known diagnosis of diabetes mellitus and on treatment, some being on follow up and some lost to follow up. The duration with diabetes in the patients in this study ranged from 3 months to 33yrs with median duration of 5(IQR 2, 8) years with several of the participants being newly diagnosed DM at the time of the DKA admission. This may indicate that despite having diabetes for longer duration education and DKA prevention efforts are still key and should be continuous on these patients with diabetes particularly on the triggers and symptoms of DKA. The duration of diabetes is a reflection to a study done in India that had a mean duration of 7±3years of the patients presenting with DKA (Sonwani et al., 2018).

Of note is the fact that all the participants in our study had poor glycemic control as reflected by their HbA1c of >7.5%, and hence were at risk for both acute and chronic complications.

Therefore, continuous education remains key and utilising opportunities during clinical reviews to ensure patients know about the importance of sugar control by adhering to their medications and prevention of complications. A study in a neighboring country; Tanzania, showed similar results of a higher number (72.4%) of DKA patients being already diagnosed DM patients (Iddi et al., 2017).

The patients who knew their diabetes status in our study and were on medication, majority (62.7%) were not adherent and the reason for non-adherence by most of them was financial constraints (46.81%), this is because most of these patients were not working or not having any business that is a source of income. This is similar to another study in India that most of the patients were non-compliant because of lack of finances (Sonwani et al., 2018).

Out of the 45% of the patients who were admitted in the past year, 10% of them had previously been admitted in DKA. Most of these patients were type 1 DM patients. This indicates that the rate of readmissions of DKA are mostly in type 1 DM and noncompliance could be the main contributor. This was similar to a study done in Riyadh Saudi Arabia which showed recurrent admissions of DKA in type 1 DM over the course of 20yrs (Almalki et al., 2016).

When comparing urine and blood ketones, some of the patients had negative urine ketones but positive blood ketones, and for some 8 patients a urine sample was not able to be taken for urinalysis but the blood ketones were significantly high for diagnosis of DKA. Out of the patients that a urinalysis was done, only 46.4% of the patients would have been diagnosed of DKA based on urine ketones alone leaving out the rest. This emphasizes the importance of the utility of blood ketones on the diagnosis of DKA rather than relying on urine ketones alone. The ability to diagnose DKA was significantly improved by use of blood ketones and this suggested that there are several cases of DKA that would have been missed because urine ketones were absent in some patients and others not significant enough (not ≥ +2) to make a diagnosis of DKA, yet they had significant blood ketones to make the diagnosis of DKA.

A significant amount of patients (63.3%) presented with leucocytosis but most of them did not have overt infection. The explanation being; most patients with DKA have leucocytosis with a left shift. This is due to dehydration and stress response to ketonemia and hyper-glycaemia by hypercortisolemia and increased catecholamine secretion and does not necessarily suggest infection. However, a white cell count greater than 25,000/μl warrants a comprehensive search for infection. This finding reflects a study done in Nairobi, Kenya in KNH (Mbugua et al., 2005) where he noted that over (65%) of patients had leucocytosis with most (55%) not having an overt infection.

Infection in this study was supported by procalcitonin (PCT) a biomarker specific for bacterial infections, C-reactive protein (CRP), CXR for pneumonia and PTB; Urinalysis for UTI and some blood cultures depending on the foci. Of the PCT done, (67.9%) was high and supported an active infection in the clinical context while (32.1%) was normal, this was done on the basis of high index of suspicion of infection and therefore it was not done on every patient.

Despite (62.5%) of the patients being known DM and on follow-up, all (100%) of the patients had a high HbA1C at the time of presentation, this indicates that all our patients had poor glycaemic control despite most of them being on follow-up in the clinics. This indicates the need of emphasis on adherence and continuous education on emphasis of glycaemic control to avoid both acute and chronic complications. This was comparable to the study done in Nairobi (KNH) that over 90% of the patients had a high HbA1C >8% (Mbugua et al., 2005).

Of all the patients that were enrolled in the study (55%) of the patients presented with acute kidney injury (AKI), signified by high creatinine and high urea levels, this was mostly due to hypovolaemia due to glucose induced osmotic polyuria and emesis since almost all patients were presenting in a dehydrated state leading to a pre-renal AKI. This was almost similar to a study on incidence and characteristics of AKI in severe DKA that recorded (50%) of patients presenting with AKI on admission (Orban, Maizière, Ghaddab, Van Obberghen, & Ichai, 2014). At the time of presentation (58%) had normal potassium levels, (12%) having low potassium and (30%) presenting with high potassium. This suggests that not all patients will have low potassium levels at the time of presentation but would still need supplementation of potassium during management because of the shift with insulin.

### Outcomes of DKA in patients seen at MTRH

The outcomes of DKA in this study were resolution of DKA, length of hospital stay and death.

The days taken before the patients were out of DKA (DKA resolution) ranged from 1 to 5 days with a median period of 3 (IQR 2, 3) days. Most of the patients remained with high sugars longer signifying that it was hard controlling their sugars while in the ward early enough and this contributed to their longer stay in the hospital. Switching of insulin was an issue that would need more education on how to go about it by following the protocol to avoid prolonged period of DKA and hyperglycaemia; also close monitoring of these patients was concerning, leading to prolonged time of resolution. This signified that DKA management was not well done since some of the patients were still having high blood ketones in day 5 resulting in them having a longer hospital stay.

The results of DKA resolution differ from a study done in Thailand whose median time of DKA resolution was 8.5hrs (Thewjitcharoen et al., 2019), another study in Melbourne had a median time of resolution of DKA of 11hrs and a median length of hospital stay being 3 days (Lee, Calder, Santamaria, & MacIsaac, 2018), the study in KNH recorded a median time of resolution of DKA to be 59hrs (2.5 days) and a study in Ethiopia recorded the mean length of hospital stay as 4.6 days (Abegaz et al., 2018).

This showed that our patients were taking longer to come out of DKA; which requires a closer audit of the management process and other possible contributing factors. The reasons for the difference in the outcome of resolution of DKA being shorter in the study by Thewjitcharoen et al was that it was done in Thailand a more developed country and also because the patients were seen in a specialised endocrine centre. Most of the patients (54.2%) were treated by endocrinologists and (45.8%) were treated by general internists or on-call physicians; (45.7%) of the patients had mild DKA, (22.3%) had moderate DKA and (31.9%) had severe DKA (Thewjitcharoen et al., 2019).

The duration of inpatient follow-up ranged from 3 hours to 10 days with a median follow-up period of 6 (IQR 5, 7) days, this was because one of the patient died within 3 hours of arrival to the hospital. The length of hospital stay was divided from day 1-5 accounting for (44.1%) and day 6-10 accounting for (55.9%). This depicted a longer length of hospital stay in most of the patients and was almost similar to a study done in Ethiopia that had a mean length of hospital stay of 4.64 (±2.802) days with about (20.4%) of patients having a longer stay of >7days (Mekonnen et al., 2022). This shows the burden that DKA puts on patients; a longer hospital stay resulting in more bills and thus the need for avoidance of this complication.

A sub-analysis done on the association between length of hospital stay and patient’s characteristics revealed a statistical significance of having infection and a history of being admitted in the last 1 year attributing to longer length of hospital stay with an OR of 2.49 and 5.74 respectively. This means that those that had infection were 2.49 times more likely to stay in the hospital longer than those who did not have infection. Those who had been admitted within the last year were 5.74 times more likely to stay in the hospital longer than those who did not have any admission within the last year. This could be attributed to the fact that those that have had several admissions are more likely to have poor glycaemic control and stay longer while those with infections staying longer because infections worsen glycemic control.

The severity of DKA, (patients with very low PH and low HCO3), rebound hyperglycaemia and infections contributed the most to longer hospital stay of these patients and this was similar to the study in Ethiopia where the longer duration of hospital stay was attributed to almost the same factors. (Abegaz et al., 2018).

We had a (9.2%) in hospital mortality in our study with (90.8%) of patients getting out of DKA and being discharged. All 11 patients who died were not out of DKA at the time of death; most of them were new onset diagnosis of diabetes mellitus with infections/sepsis and acute kidney injury as the major comorbid that could have contributed to death. Eight (72.7%) died within the 1st day of follow-up within 48hrs from admission, most of them were referrals and had reduced level of consciousness, hypotension, hypokalaemia and severe metabolic acidosis. This gives median follow-up period for those who died to be 1 (IQR 0.25, 2) days and 6 (IQR 5, 7) days for those who were discharged alive. This is a very significant finding that most patients died on the first day of diagnosis of DKA; it could indicate that the patients were coming in severely ill and in delayed presentation with altered level of consciousness, or with comorbid like infections and with other complications like hypokalaemia that contributed to their mortality. It also questions if appropriate management according to protocol was implemented in time because reviews by the registrars were delayed and close monitoring was not efficiently done. This is significant for future research. Most of the patients were also managed in the general medical wards and close monitoring was wanting because of the many number of patients in the wards as a referral facility with few nurses yet these patients ought to be managed in the ICU or HDU in an ideal setting. We can compare our mortality to be equal to Ethiopia’s mortality rate back in 2015 that was at 9.8%. But a more recent study in Ethiopia by Mekonnen et al had a low mortality of (4.4%) but it also had a larger percentage of patients (75.5%) with mild DKA and few severe DKA (Mekonnen et al., 2022).

A sub analysis done on the association between the outcome of death and patient’s characteristics, revealed that those who were newly diagnosed DM were 5.19 times more likely to die with a p value of 0.02. This is closely related to a study in Iran by Razavi et al that demonstrated that mortality appeared to be greatest among patients at first presentation mostly in newly diagnosed T1DM patients (Razavi, 2010). This can be explained by the acute stress response in the newly diagnosed that can be overwhelming.

For comparison, the mortality rate in the KNH study had (29.8%) of their subjects dying within 48hrs or admission (Mbugua et al., 2005). A follow-up study on the prognostic factors in patients hospitalised with DKA in KNH showed that altered level of consciousness was a major predictor for mortality (Otieno, Kayima, Mbugua, Amayo, & McLigeyo, 2010). A retrospective study done in Ethiopia between 2015 and 2017 concluded that DKA contributed to (12%) in-hospital mortality (Taye, Bacha, Taye, Bule, & Tefera, 2021).

In a recent study, the overall mortality rate of DKA is (0.2-2.0%) with persons at the highest end of the range residing in developing countries (Hamdy, 2019), therefore the mortality rate we had was higher than the expected rate and improvement is needed to reduce this outcome. In order to have a better outcome, clinicians should observe the four pillars of treatment of DKA promptly; correction of fluid loss with intravenous fluids, correction of hyperglycaemia with insulin, correction of electrolyte disturbances particularly potassium and correction of acid base balance with recognition and treatment of precipitating causes. Close monitoring of patient’s condition by regular clinical and laboratory data and the use of management protocols help ensure better outcomes. Therefore, these patients should ideally be managed in an intensive care unit (ICU) or high dependency unit (HDU) setting where close monitoring is done.

## Conclusion

This study has demonstrated that DKA occurs in undiagnosed DM patients and affects both T1DM and T2DM patients.

Some of the identified DKA precipitants in the study are preventable.

Most of the patients were presenting in a dehydrated state, hence the importance of using blood ketones as a point of care measurement for the diagnosis of DKA at presentation rather than depending on urinary ketones.

Finally, the study has shown that DKA is associated with significant hospital stay and mortality in MTRH.

## Recommendations

We would recommend increased public education on symptoms and signs of diabetes mellitus to improve uptake of screening and early presentation to hospital. Once a diagnosis of DM has been made on either type of DM, screening for DKA should be done and timely management strategies provided according to protocol.

Since missed doses was an important precipitant of DKA, we would recommend availability and affordability of drugs and that clinicians should emphasize on adherence during prescription of these drugs.

We would recommend use of blood ketones as a point of care measurement for the diagnosis of DKA since it significantly improved the diagnosis of DKA. For the availability of these kits in the Emergency Department and the DM clinics. With a high index of suspicion, patients presenting with hyperglycaemia irrespective of the type of DM should be subjected to blood ketones measurement in order not to miss out DKA.

Lastly we recommend further research that can be done on the audit of management of DKA since it was not fully captured in the study, to find out if protocols are well observed/adhered to and a follow up study if outcomes will improve. We also need more endocrinologists to help in the management of these patients.

## Data Availability

All data has been availed in the manuscript

### Appendices

#### Appendix 1: QUESTIONNAIRE

STUDY INVESTIGATOR............................................

DATE OF ASSESSMENT............................................

STUDY NO................................................................

DEMOGRAPHIC/PATIENT DETAILS

~~~
1) Age () yrs
2) Gender: 1. Male () 2 Female ()
3) Marital status: Single () Married ()
 Separated () Widowed ()
4) Education Level: None () Primary ()
 Secondary () Tertiary ()
5) Occupation/ Employment: Unemployed ()
 Formally employed ()
 Self employed ()
 Student ()
6) Living arrangement: Alone ()
 With others ()
~~~

MEDICAL HISTORY

~~~
7) Diabetes Mellitus type: Type 1 () Type 2 ()
8) Age at diagnosis of diabetes ........
9) Duration of diabetes followup: < 6months () 6m-1 yr ()
 1-5yrs () >5yrs ()
10) Current medication: (Drugs and dosages)
Oral anti diabetic agents ........................................................
 Insulin .......................................................................................
 Both ..........................................................................................
 Others....................................................................
 ............................................................................
11) Are you compliant to the above treatment: Yes....................or No.....................
If No, please explain why
..............................................................................................................................................................................................
................................................................................................................................................................................................
12) Have you had any admissions in the last 12months? If yes it was due to
..........................................................................................................
..........................................................................................................
........
13) Precipitants: Infection (YES/ NO):
 Pneumonia-cough, chestpain ()
 Urinary tract infection; dysuria, polyuria ()
 Vaginal candidiasis; vulval itchiness and whitish discharge ()
 Pulmonary tuberculosis: cough, weight loss, night sweats ()
 New onset of DM ()
 Dosing issues: Missed doses (), Underdosing ()
 Injection technique: lipodystrophy (), wrong injection sites ()
 Insulin storage: storage in warm places ()
 Storage in water ()
 Storage in freezer ()
 Prolonged use of open vial >1 month ()
 Prescribed drugs eg thiazides, steroids ()
 Stresses: Myocardial Infarction: severe chest pain ()
 Cerebrovascular accident: focal weakness ()
14) Clinical Presentations: History
 Polyuria ()
 Polydypsia ()
 Polyphagia ()
 Nausea and vomiting ()
 Abdominal pain ()
 Weight loss ()
 Physical Examination:
 Vitals: BP (), Temp(), HR(), RR()
 Dehydration ()
 Kussmaul Breathing ()
 Level of consciousness ()
~~~

Clinical tests:

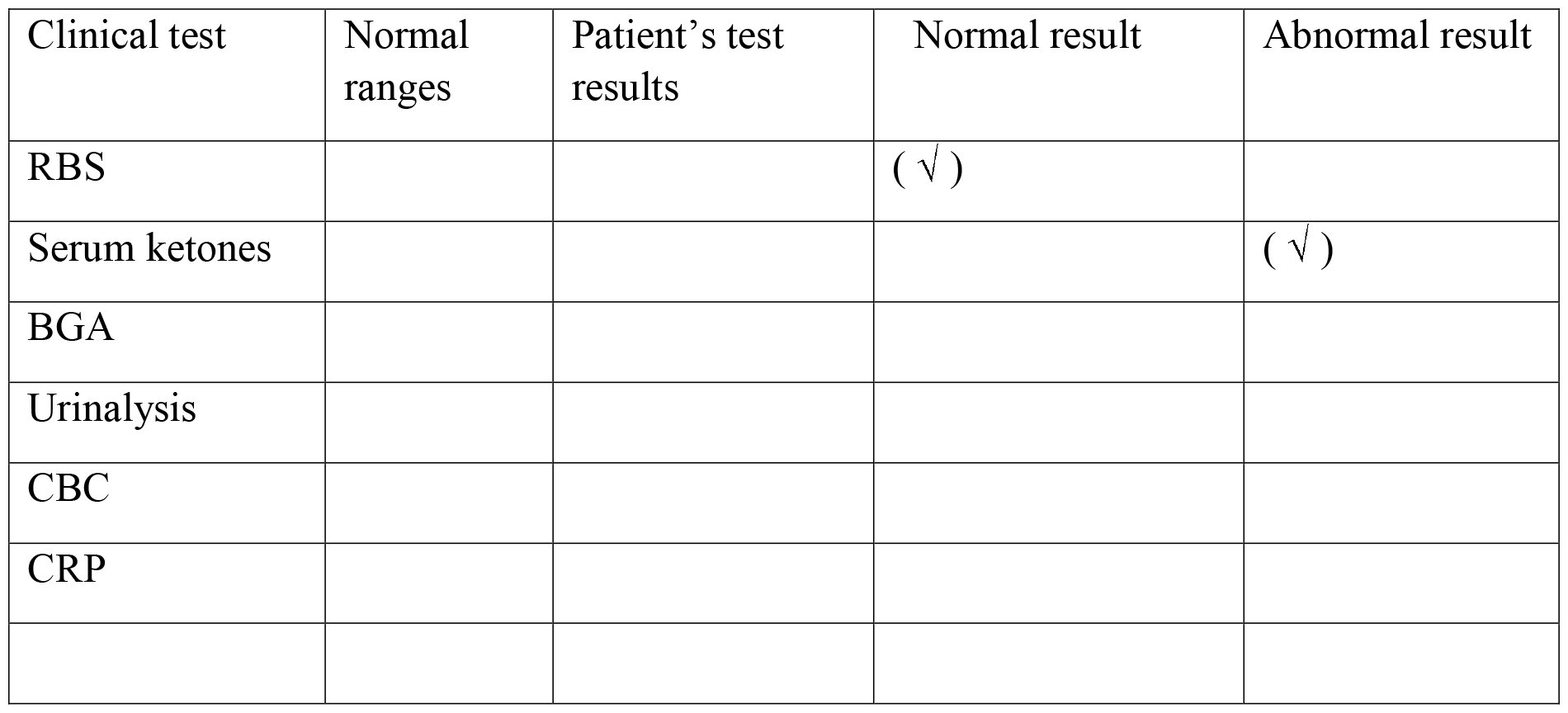

| Clinical test | Normal ranges | Patient's test results | Normal result | Abnormal result |
| --- | --- | --- | --- | --- |
| RBS |  |  | ( √ ) |  |
| Serum ketones |  |  |  | ( √ ) |
| BGA |  |  |  |  |
| Urinalysis |  |  |  |  |
| CBC |  |  |  |  |
| CRP |  |  |  |  |

OUTCOMES

Resolution of DKA: Resolution of DKA defined according to definition; arterial/venous Ph >7.3 and/or venous bicarbonate >18mmol/l yes............ no.................

What date and time was the resolution of DKA identified .........................

Length of hospital stay: mean days of hospital stay ................minimum days .................. maximum days .............

Death Yes .......... No....

#### Appendix 2: INFORMED CONSENT (ENGLISH)

##### Introduction

My name is Dr. Clemence Msagha, a postgraduate student of internal medicine at Moi Teaching and Referral Hospital. I am the principal investigator(PI) in a study on the precipitating factors, clinical presentations and outcomes of DKA in MTRH. I would like to request you to participate in this study. This is a consent form that gives you information about this study.

You are encouraged to ask for clarification at any point where information is not clear. After reading and understanding the information provided, you will be required to sign the informed consent form.

Parents/ guardians/ next of kin will sign on behalf of patients who are less than 18yrs of age or those that are unable to sign the consent due to disease severity (reduced level of consciousness). Participants less than 18yrs of age will be required to sign assent forms prior to inclusion into the study.

Participation is voluntary and one has a right to withdraw at any point of time and there will be no monetary incentives. One will not be victimised if they refuse to participate in the study.

##### Purpose of the study

The study is being done on people living with diabetes either type 1 or type 2. DKA is a complication of DM. I will be looking at the different precipitating factors that different patients will present with, the clinical presentations and the outcomes of patients seen in MTRH.

This research will be of benefit because you will participate in increasing information on the common precipitating factors, presentation and outcomes of DKA in our setup and help in improving the early diagnosis and management of DKA in MTRH.

##### Procedures of the study

If you decide to join this study you will be required to fill in a questionnaire, this will contain your basic details: age, sex, occupation; health assessment and risk assessment questionnaire. A brief medical history will be taken including your type of diabetes, duration of diabetes, current medication, any of the precipitating factors that will be in the questionnaire, and a physical examination done and samples taken.

There will be confidentiality in all the information given in that data given will be kept locked in a secure location at all times and only those who are directly involved in the research will have access to them. We will be using initials or identification number and not full names. Any publication of this study will not use your name or identify you personally. Efforts will be made to keep your personal information confidential. We cannot guarantee absolute confidentiality.

Your personal information may be disclosed if required by law.

##### Your consent

I have been adequately informed that I am being recruited in a study to find out the precipitating factors, presentations and outcomes of DKA in DM patients. The investigator has also informed me that my participation in this study is voluntary and will not exclude me from routine care even if I were to opt out. She has also informed me that I’ll not be required to pay for the tests done for the purposes of this study.

I ................................................................................ have read the information above and it has been explained to me by Dr Clemence Mwahe Msagha or a research assistant. I voluntarily agree to participate in the study ‘Diabetic Ketoacidosis among people living with diabetes seen at Moi Teaching and Referral Hospital’

**CONSENT (Parents/Guardian/Next of kin/Participants >18yrs of age)**

**SIGNED/THUMBPRINT................................................................**

**NAME: ............................................................................................**.

**DATE............................................................................................**....

**ASSENT FORM (Participants <18yrs of age)**

The researcher has explained to me about the study

I understand the researcher will ask questions about me

I understand they will take a little blood from me

They need this blood to find out if I have Diabetic ketoacidosis

My parent/guardian has agreed that I should participate in the study, I also voluntarily agree that I should participate to answer any question asked and samples be taken.

**SINGED/THUMBPRINT................................................................**.

**NAME OF CHILD ........................................................................**.

**DATE................................................................................................**

**INVESTIGATOR OR RESEARCH ASSISTANT STATEMENT**

I have provided an explanation of the purpose of this study to the participants.

**SIGNED....................................................................................**

**DATE........................................................................................**.

For further clarification you may contact:

Dr Clemence Msagha 0724978121

